# AI coupled to pharmacometric modelling to tailor malaria and tuberculosis treatment in Africa

**DOI:** 10.1101/2024.11.07.24316884

**Authors:** Gemma Turon, Mwila Mulubwa, Anna Montaner, Mathew Njoroge, Kelly Chibale, Miquel Duran-Frigola

## Abstract

Africa’s vast genetic diversity poses challenges for optimising drug treatments in the continent, which is exacerbated by the fact that drug discovery and development efforts have historically been performed outside Africa. This has led to suboptimal therapeutic outcomes in African populations and overall scarcity of relevant pharmacogenetic data, including characteristic genotypes as well as drugs prescribed in the continent to treat infectious diseases. Here, we propose a general approach to identify drug-gene pairs with potential pharmacogenetic interest. Furthermore, we delve deeper into the analysis of malaria and tuberculosis therapies, many of which remain uncharacterised from a pharmacogenetic perspective. Our pipeline leverages artificial intelligence and the latest advances in knowledge embedding techniques to exploit currently available biomedical data and subsequently prioritise pharmacogenes for each drug. Predicted pharmacogenes are then incorporated into pharmacometric modelling to hypothesise which ones might be of clinical interest, and which dose adjustments could be made to provide better treatment outcomes for the African population.

## Introduction

The African continent is the most genetically diverse, displaying a wide range of genetic variants that may influence patient responses to drug treatment. Such diversity presents challenges and opportunities in the field of pharmacogenomics (PGx), which seeks to adjust drug therapies based on genetic information, maximising treatment efficacy and minimising adverse effects. Unfortunately, widespread application of PGx in Africa is hindered by the scarcity of available data, particularly in the context of drugs for endemic infectious diseases and genetic variants prevalent among African populations. As a result, comprehensive PGx datasets and subsequent data analysis studies of relevance to Africa are virtually non-existent.

In other stages of drug discovery and development, computational approaches, including artificial intelligence (AI) and machine learning (ML) have demonstrated significant value, providing, among many others, target candidates, predicted inhibitors for these targets, hit-to-lead optimization suggestions, etc. However, streamlined application of computational procedures to PGx is lagging behind^1^, with few anecdotal examples available in the literature^2,3^, and a lack of well-established tools freely available to the scientific community, even for the most seemingly simple tasks such as predicting which ‘pharmacogenes’ might be relevant to a given drug, much like other tools predict protein-ligand interactions. Arguably, while conceptually similar, the PGx task is significantly more challenging, with 2-3 orders of magnitude less data available in the public domain, and only related to a fraction of the approved drugs. Additionally, PGx relationships are not immediately modellable using physics-based approaches as is the case with molecular docking for protein-ligand interactions, making it difficult to apply computational techniques when previous evidence or training data is scarce.

In recent years, AI and ML methods have evolved to operate in low-data scenarios, increasing their capacity to integrate data from multiple sources and apply it to the task of interest. Indeed, there is a vast amount of biomedical data related to human genes and approved drugs that could potentially be leveraged to anticipate PGx interactions. For example, known physical interactions between drugs and cytochrome enzymes may inform their PGx profiles, and drugs with similar indications and chemical characteristics may tend to elicit PGx interactions with the same pharmacogenes. From an ML/AI perspective, these connections can be systematically explored through large-scale integration of biomedical knowledge, followed by a mapping between known PGx interactions and this somewhat orthogonal and more abundant information. Moreover, the advent of large language models (LLMs), which increasingly demonstrate proficiency in biomedical and pharmaceutical domains, is opening a whole new range of opportunities in PGx that remain largely unexplored.

Recently, the Project Africa GRADIENT (Genomic Research Approach for Diversity and Optimising Therapeutics) initiative was launched with the goal of exploring how genetic variability across the African continent might influence current therapies, particularly for malaria and tuberculosis (TB), which continue to impose a high burden in the region^4^. This initiative includes our project focussed on exploring the potential of ML/AI in PGx, given the critical need for ‘hypothesis generation’ in this area and the relatively low capacity to collect data in the short term. This makes computational predictions provided by ML/AI particularly impactful and enabling. Current AI strategies focus on mining the existing literature^5^. Instead, we set out to prioritise new putative pharmacogenes for over thirty malaria and TB drugs prescribed in Africa, overlaying information about the abundance of genetic variants found in those genes among the population. We then used these prioritised genes to adjust and improve the pharmacometric models related to each of the drugs, demonstrating for the first time how ML/AI PGx predictions can be systematically integrated into physiologically-based pharmacokinetics (PBPK) and nonlinear mixed-effects (NLME) models^6^. Here, we show how this genuinely novel computational approach could aid the adjustment of dosing regimens for malaria and tuberculosis drugs in Africa. We demonstrate the cases of artemether and rifampicin end-to-end and discuss how the methodology could be applied to other drugs and diseases areas, both in Africa and elsewhere.

## Results

### Scarce PGx information in Africa

As a ground truth dataset, we curated PharmGKB^7^, the largest publicly available database of pharmacogenetic drug-gene interactions, and obtained a list of 1,111 drugs for which there is, at least, one PGx association reported. A PGx association is characterised by a chemical-gene-variant triplet that has been reported in the scientific literature or is part of a recommendation report, for example, from the CPIC consortium. Importantly, one pair of chemical-gene can have multiple annotations if several variants from the same gene have been reported to interact with the given chemical. In addition, the association can influence the pharmacokinetics (PK) or pharmacodynamics (PD) of the drug, including efficacy, dosage and toxicity considerations. To provide an overview of the data available in PharmGKB, at this stage of the analysis we consider both PK/PD effects. Of the annotated drugs, we found that only 14.2% of them are indicated for communicable diseases (Figure 1A). Moreover, infectious disease-related drugs have on average, less annotations per drug when compared to non-communicable diseases (Figure 1B). These data highlight the need to increase PGx studies in infectious diseases.

**Figure 1.**
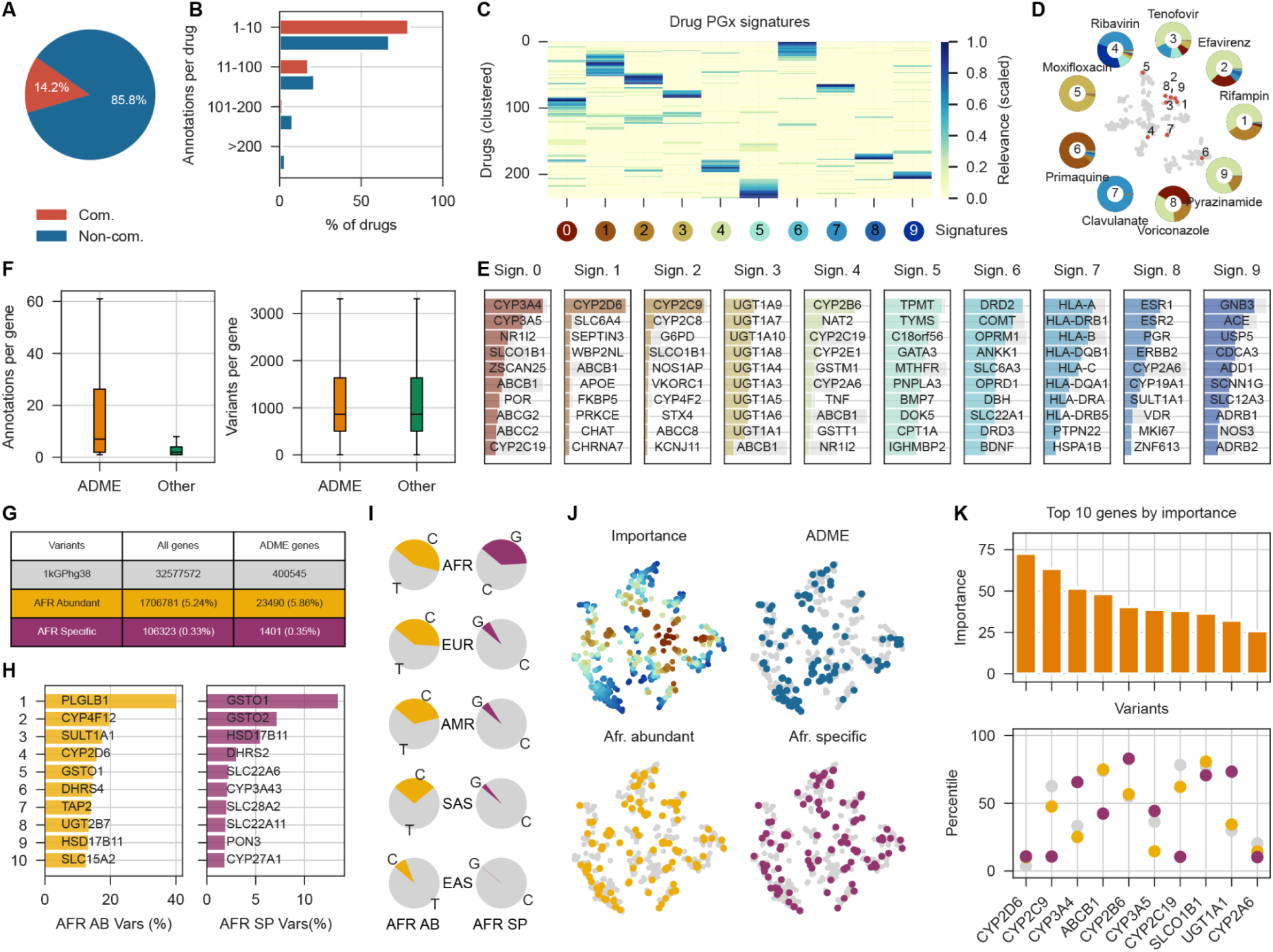
Overview of PharmGKB data and African genomic variants. **A.** Percentage of drugs for communicable vs non-communicable diseases with at least one significant pharmacogenetic association in PharmGKB. **B.** Distribution of communicable vs non-communicable drugs according to their number of PharmGKB annotations. **C.** Clustering of drugs according to their PGx signature. **D.** T-SNE clustering of drugs according to their PGx Signature. Examples of infectious disease drugs belonging to different signature groups are highlighted. **E.** Number of PharmGKB annotations and number of genomic variants in ADME vs Non-ADME genes. **F.** Top ten genes in each of the PGx gene signatures. **G.** Percentage of African Abundant and African Specific variants in the 1kGPhg38 whole genome annotation and specifically in ADME genes. **H.** Top ten ADME genes by number of African Abundant and African Specific variants. **I.** Example of an African abundant variant distribution across populations corresponding to rs7255816 (gene: CYP4F12) and example of an African specific distribution across populations corresponding to rs680055 (gene: CYP3A43) (AFR: Africa, EUR: Europe, AMR: America, SAS: South-Asia, EAS: East-Asia). **J.** T-SNE of PharmGKB-annotated genes represented by signature association. Overall importance in signature analysis is shown in the top left, ADME genes are highlighted in the top right, and top 10% of genes according to African abundant and African specific variants are shown in the bottom. **K.** Top ten genes selected by their importance to PGx signature and representation of the African Abundant (yellow) and African Specific (purple) variants they harbour.

To gain a global understanding of PharmGKB data, we then decomposed the matrix of drug-gene interactions into two much more compact drug-signature and signature-gene matrices, where signatures represent sets of pharmacogenes that tend to co-occur across drugs. For this part of the analysis, only drugs with at least 5 annotated pharmacogenes were considered. In Figure 1C, at a high level, it can be seen how drugs can be partitioned into 10 pharmacogene signatures, with some drugs being very specific to one signature (e.g. clavulanate and moxifloxacin) and others showing a more varied profile (e.g. ribavirin and voriconazole) (Figure 1D). Interestingly, seemingly unrelated drugs such as tenofovir, rifampicin and pyrazinamide, shared association with the same signature (Figure 1D) defined by pharmacogenes such as CYP2B6 and NAT2 (Figure 1E). Of note, some signatures were specific to one or a few genes (e.g. signature 0: CYP3A4 and CYP3A5, which co-occur as expected; signature 1: CYP2D6, signature 2: CYP2C9), while others encompassed gene families (signature 3: UGTs, signature 7: HLAs).

In addition, we were interested in understanding the distribution of PharmGKB annotations by gene, in particular comparing ADME vs non-ADME genes. We used a list of 283 ADME genes curated by the PharmADME working group, defined by their involvement in the administration, distribution, metabolism or excretion of drugs^8^. As such, ADME genes are expected to be more frequently associated with PGx effects. Indeed, we observed that in our curated data ADME genes harbour a much higher proportion of significant annotations in PharmGKB when compared to non-ADME genes (Figure 1E).

In parallel to the curation of PharmGKB data, we also focused on the identification of genes that will be more relevant to downstream applications of our study because they carry variants that are prevalent in Africa. To that end, we used the 1000 Genomes Project data^9^ realigned to GRC38 (1kGPhg38), which contains whole genome sequencing (WGS) and deep whole exome sequencing (WES) of 2,504 individuals from 26 different populations. The populations are grouped into the following categories: Africa (AFR), Europe (EUR), America (AMR), South Asia (SAS), East Asia (EAS). First, we analysed the distribution of variants between ADME and non-ADME genes, demonstrating that, despite ADME gene variants being annotated much more frequently in PharmGKB, both ADME and non-ADME genes harbour a similar number of genetic variants (Figure 1F). Next, we set out to classify the variants as non-African (i.e. variants that are present in less than 20% of the African samples), Africa-abundant (i.e. variants that are present at a higher proportion in African populations, but are also common to others; AFR-abundant) or Africa-specific (i.e. variants that, on top of being abundant, are present at a much higher proportion with respect to any other ancestry; AFR-specific). In sum, an AFR-abundant variant can be either private or shared with other populations, while an AFR-specific variant is necessarily private to African populations. To that end, we first determined a cut-off for African abundance as an Allele Frequency (AF) ≥ 0.20, as suggested in an earlier study^10^. This rendered a total of 1,734,675 AFR-abundant variants, which represented the top 5.5% of the data, since the AF distribution of African variants 1kGPhg38 annotated dataset is mainly found at AF < 0.1 (Figure S1A,B). Subsequently, we sought to determine which of the AFR-abundant variants were actually AFR-specific by using an 8x enrichment factor of AF in AFR vs other populations. This threshold was determined by observing the distribution of AFR-specific variants within our dataset as defined by Fedorova et al^10^. The distribution showed a peak at approximately 12x and a minimum at around ∼8x (Figure S1C). As a result, a total of 177,364 AFR-specific variants were obtained. These represent a 0.60% of the 29,459,588 1kGPhg38 dataset and a 10.22% of the AFR-abundant variants. A total of 2,285 (1.29%) of these AFR-specific variants were located within 186 of the ADME genes. Overall, ADME genes maintained the proportion of AFR-abundant and AFR-specific variants in ADME vs non-ADME genes (Figure 1G). Among the genes harbouring more AFR-abundant and AFR-specific variants, we find members of the CYP450 family and numerous transporters (Figure 1H). An example of the AF distribution across populations of an AFR-abundant and AFR-specific variant is showcased in Figure 1I. The distribution of AFR-abundant and AFR-specific variants annotated in PharmGKB is maintained between ADME and non-ADME genes (Figure S2).

Finally, we plotted the PharmGKB-annotated genes according to their signature association (Figure 1J). The top 10 genes important to the PGx signatures are relevant ADME genes, including several CYP450 family members and transporters (Figure 1K). Interestingly, genes such as CYP3A4, CYP2B6 and UGT1A1 harbour a relatively high proportion of AFR-specific variants.

### An AI-based methodology to prioritise pharmacogenes

Next, we set out to develop an ML/AI method to predict drug-pharmacogene pairs, using PharmGKB as a reference dataset. Given the scarce PGx information available, we devised a strategy that leverages orthogonal information available for drugs and genes to assist in the training procedure. Previously, in tasks such as drug activity prediction, we showed that incorporating publicly available data from chemoinformatics and bioinformatics resources can improve the predictive power of ML models, especially when few training data points are available^11^. Here, we primarily used a biomedical knowledge graph that collects drug and gene information of multiple types, including drug-gene interactions, gene expression profiles in cells, drug side effects, protein function and cellular localization, etc. From this knowledge graph, we selected types of biomedical relationships that we thought made sense in the context of PGx, for example, protein abundances in tissues, protein-protein interactions, participation of proteins in pathways, and drug targets and metabolic genes (Tables S1 and S2). This information was retrieved in ‘embedded’ form from the Bioteque^12^, i.e. in 128-dimensional vectors assigned to each drug and each gene in our dataset. In brief, for each type of relationship and data source (e.g. protein-protein interactions from the STRING database^13^) in the knowledge graph, we obtained an embedding vector for each entity (gene, in this case), such that similar vectors correspond to proximal nodes in the original network. In addition to knowledge graph embedding vectors, we also obtained sequence embeddings for genes^14^ (i.e. learned numerical representations of protein sequences such that similar embeddings correspond to similar sequences), as well as multiple descriptors for the drugs. As drug descriptors, we used a representative repertoire of tools, including 2D chemical fingerprints, arrays of physicochemical properties^15^, bioactivity profiles, and ADME calculations^16^ (Table S3). Collectively, we assembled, for each drug and gene in our dataset, a compendium of numerical arrays that captured the knowledge available for them in a vectorial form that is amenable to downstream ML tasks (Figure 2A).

**Figure 2.**
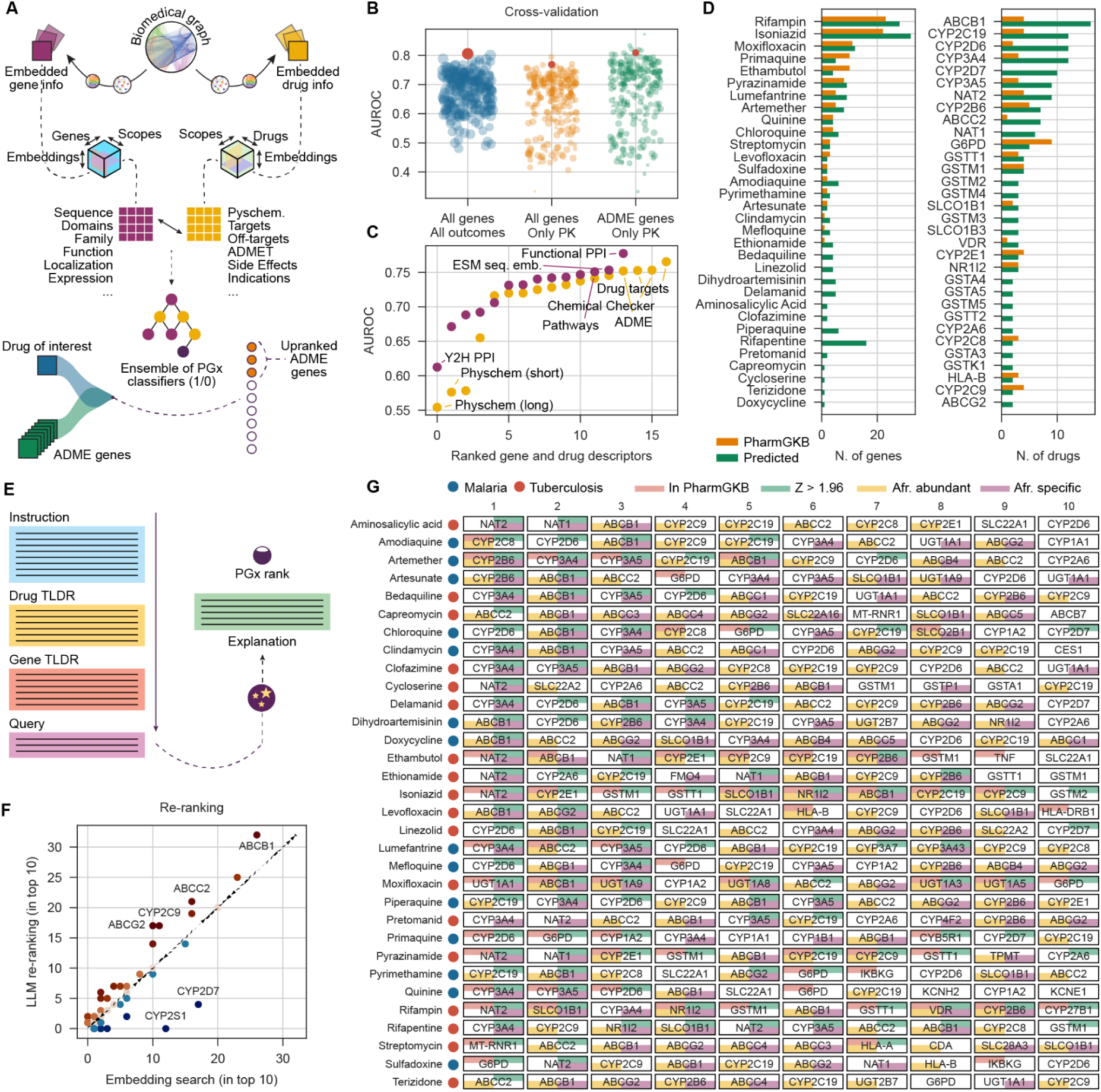
AI pipeline for drug-pharmacogene prioritisation in malaria and tuberculosis. **A.** Graphical representation of the pipeline to extract gene and drug embeddings for the training of the PGx classifier. **B.** Cross-validation performance measured by the Area Under the Curve (AUROC) of the different PGx classifiers and the performance of each of the three ensembles (red dot). **C.** Top quartile performance (AUROC) of classifiers using different individual drug and gene descriptors. **D.** Number of genes associated with each of the selected drugs for the study according to PharmGKB vs predicted by the PGx classifier, and number of selected drugs associated with the top genes according to the PGx classifier vs the number of annotations in PharmGKB. **E.** Graphical representation of the LLM interrogation for the selection of the top ten PGx associations per drug. **F.** Genes in the top ten most important PGx associations to the drugs of study according to the LLM vs the initial PGx classifier. **G.** Top ten genes with predicted PGx association per drug (z-score >1.96, green) and annotated by their association in PharmGKB (orange). In yellow, we show genes that have at least 50 AFR-abundant variants and, in purple, genes that have at least one AFR-specific variant.

While the collected orthogonal information above is not explicitly related to PGx, it constitutes a good starting point for a supervised ML approach whereby a drug-gene pair predictor is trained based on their known drug-gene associations in PharmGKB (outcome variable; *y*) and the drug/gene embeddings or vectors (features; *X*). More precisely, we formulated the problem as three binary classification tasks, corresponding to three training sets of decreasing size, namely all drug-gene pairs in PharmGKB with any type of PGx association, all drug-gene pairs with a PK association, and drug-ADME gene pairs with a PK association (Figure 2B). For each binary classification task, we defined as positive (1) the known drug-pharmacogene associations, and we randomly sampled a set of 10x unlabelled drug-pharmacogene pairs to be used as negatives (0), preserving the relative frequency of drugs and genes observed in PharmGKB. We then trained an ensemble of models for each task in a fully automated manner, each member of the ensemble corresponding to a model trained with a certain type of drug and gene features (e.g. drug side effects and protein-protein interactions) (Materials and Methods). In a cross-validation procedure, we found that aggregating results across the ensemble consistently yielded high performance (Figure 2B), with AUROC scores around 0.8 for all three binary classification tasks. Of note, we found that biologically-informative representation for drugs such as drug target profiles, Chemical Checker bioactivity signatures^17,18^, and predicted ADME properties (including, among others, interactions with CYPs), were most powerful predictors, whereas purely physicochemical descriptors as commonly used in QSAR modelling yielded least performant models. Similarly, functional protein-protein interactions (PPIs), protein sequence embeddings (which are known to capture molecular function and structural features), and protein annotated pathways were most predictive, while physical (not necessarily functional) PPI interactions detected *in vitro* in a yeast-two hybrid (Y2H) assay were the least informative in this setting (Figure 2C).

We then carried out an exhaustive prediction experiment evaluating all pairwise combinations of drugs and genes. To simplify the analysis, we empirically derived a consensus Z-score averaged across the three models. In Figure 2D, it can be seen that for a majority of the drugs of our interest (i.e. malaria and TB drugs), we could find significant predictions, including drugs such as rifapentine for which no information was available in PharmGKB. Likewise, ADME genes such as NAT1 or ABCC2, which had no or little annotation in PharmGKB, appeared to be associated with a relatively high number of malaria and TB drugs.

In modern AI search algorithms, embedding-based search is often used as an initial ranking procedure that is then refined with a large language model (LLM). We therefore submitted our ranked list of pharmacogenes predicted for each drug to additional scrutiny using GPT-4, which has demonstrated competitive performance in biomedical domain settings^19^. In particular, we took the top-50 genes (ranked by Z-score) predicted for each drug, and asked GPT-4 to select 10 of them. This selection was done using controlled prompt-engineering and including prior knowledge about drugs and genes as context for the LLM (Figure 2E and Table S4). In Figure 2F, we can see that, while there was a correspondence between the top 10 genes selected in the embedding-based search and the final LLM, some genes were favoured by the LLM, including transporters such as ABCB1, ABCC2, ABCG2, and the CYP2C9 enzyme. On the other hand, CYP2S1 was deprioritised by the LLM, as well as the segregating pseudogene CYP2D7.

Overall, we obtained, for each of the 32 malaria and TB drugs of interest, a list of top 10 ADME pharmacogenes predicted by our pipeline (Figure 2F). Some of the predicted associations were already known PGx associations, such as the relationship between rifampicin and NAT2, or the association between sulfadoxine and G6PD. These are marked in red in Figure 2F. However, most of the associations have not previously been reported. For example, bedaquiline, a relatively new (2012) approved drug to multidrug resistant tuberculosis, is not annotated in PharmGKB and we predicted it to be associated with CYP3A4 and CYP3A5 (which, reassuringly, tend to co-occur in the literature^20^) as well as ABCB1, among other pharmacogenes. Indeed, ABCB1 was predicted to be associated with several drugs, which might indicate that this is a gene worth exploring for these disease areas generally. ABCB1 has also been associated with malaria severity, both through effects on drug response and through affecting the host’s interaction with *P. falciparum*^21^. Interestingly, ABCB1 harbours AFR-abundant and AFR-specific variants, as do other ABC transporters like ABCC3, ABCC4 and ABCG2, also found in our panel of predictions. To make the results of this ML/AI-based prediction available to the community, we have released a web application as specified in the Code Availability section. In this web application, we include LLM-generated summaries for the drugs and genes, and their predicted associations, with the hope that they can guide researchers into interpreting the predictions and critically assessing them.

### Integrating predicted pharmacogenes into PBPK modelling

Having prioritised 10 pharmacogenes per drug, we then asked whether these predictions could be incorporated into PBPK modelling (Figure 3A). To quantitatively assess this, we performed a comprehensive sensitivity analysis of 10 of our drugs for which we could find evidence of variable PK and drug response in African cohorts (Table S5). In the sensitivity analysis, the AUC and Cmax PK parameters were evaluated for their fold-change if the predicted genes were incorporated into the model with respect to the absence of those genes (Materials and Methods). In Figure 3B, we can see that, for all drugs, we found at least one sensitive pharmacogene, with CYP3A4, CYP3A5 and ABCB1 being frequent across drugs. Importantly, of the 23 sensitive drug-pharmacogene pairs (Figure 3B), 12 (52%) were not previously reported in PharmGKB, demonstrating the potential of our approach to quickly prioritise pharmacogenes, especially for drugs for which few or no PGx interactions are known. For amodiaquine, for example, only CYP2C8 of the 5 sensitive genes was reported in PharmGKB, while newly predicted genes such as ABCB1 and ABCG2 elicited even higher sensitivity scores. Moreover, when we randomly sampled 10 genes from the ADME list, we could not identify any sensitive drug-gene pair, demonstrating the significance of our results. Of the newly identified PBPK-sensitive drug-pharmacogene associations, 61% involved genes harbouring at least 50 AFR-abundant variants, and 87% involved genes with at least one AFR-specific variant, suggesting that our findings are worth exploring more in depth in the context of African genetic studies. Notably, >80% of the variants in our dataset corresponded to intronic variants (annotated with SNPeff^22^, see Materials and Methods), which may be particularly relevant in the context of PBPK modelling, since intronic variants tend to be associated with gene regulation. This is coherent with the PBPK software PK-Sim^®^ used for the analysis, which utilises gene expression data as a proxy for protein abundance to estimate the in vivo activity of enzymes and transporters that affect PK^23^.

**Figure 3.**
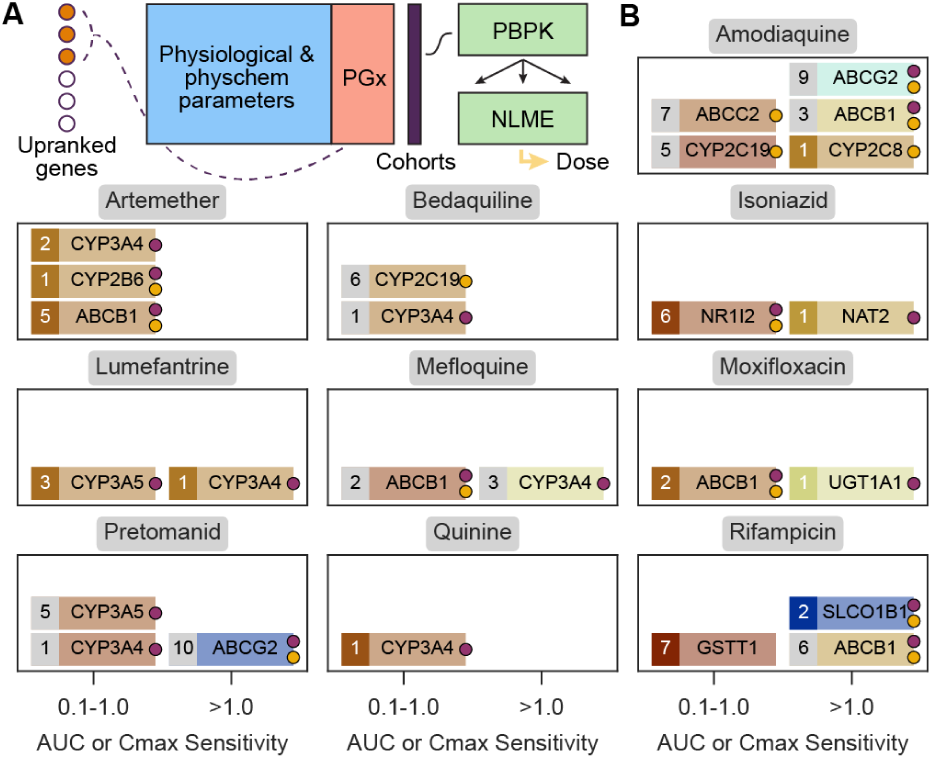
Incorporating PGx information into pharmacometric modelling. **A.** Graphical representation of the pipeline to include PGx information into PBPK and PK modelling. **B.** Sensitivity analysis by PBPK of the top 10 predicted PGx associations between selected drugs and ADME genes. Genes with a sensitivity between 0.1 and 1 inclusive are shown in the left column of each panel. Genes with sensitivity values greater than one are shown in the right column. Red-to-blue spectral scale is for the sensitivity values (low-to-high). The number indicates the rank in our predicted list. Numbers in a grey box indicate genes not reported in PharmGKB with any level of evidence for the drug. Numbers in a coloured box indicate that some degree of evidence is available in PharmGKB.

To illustrate how the pharmacometric analysis could proceed from this point, we chose the case of rifampicin and artemether (Figure 4A). For artemether, the CYP3A4, CYP2B6 and ABCB1 pharmacogenes were sensitive. While association with CYP3A4 and CYP2B6 was previously reported in PharmGKB, for ABCB1 we found further support in the literature^24^ (there was only low-level evidence in PharmGKB via an automated annotation). The simulated concentration-time profile of artemether in the presence and absence of ABCB1 transporter protein is shown in Figure 4B. ABCB1 was also found to be sensitive in the PBPK analysis of rifampicin, and has also recently been associated with rifampicin pharmacokinetics^25^. Another highly sensitive (>1) gene, SLCO1B1 is known to play a role in rifampicin PK^26^ and was considered jointly with ABCB1 for further analysis. The full PBPK models files of artemether and rifampicin are included as supplementary Table S6.

**Figure 4.**
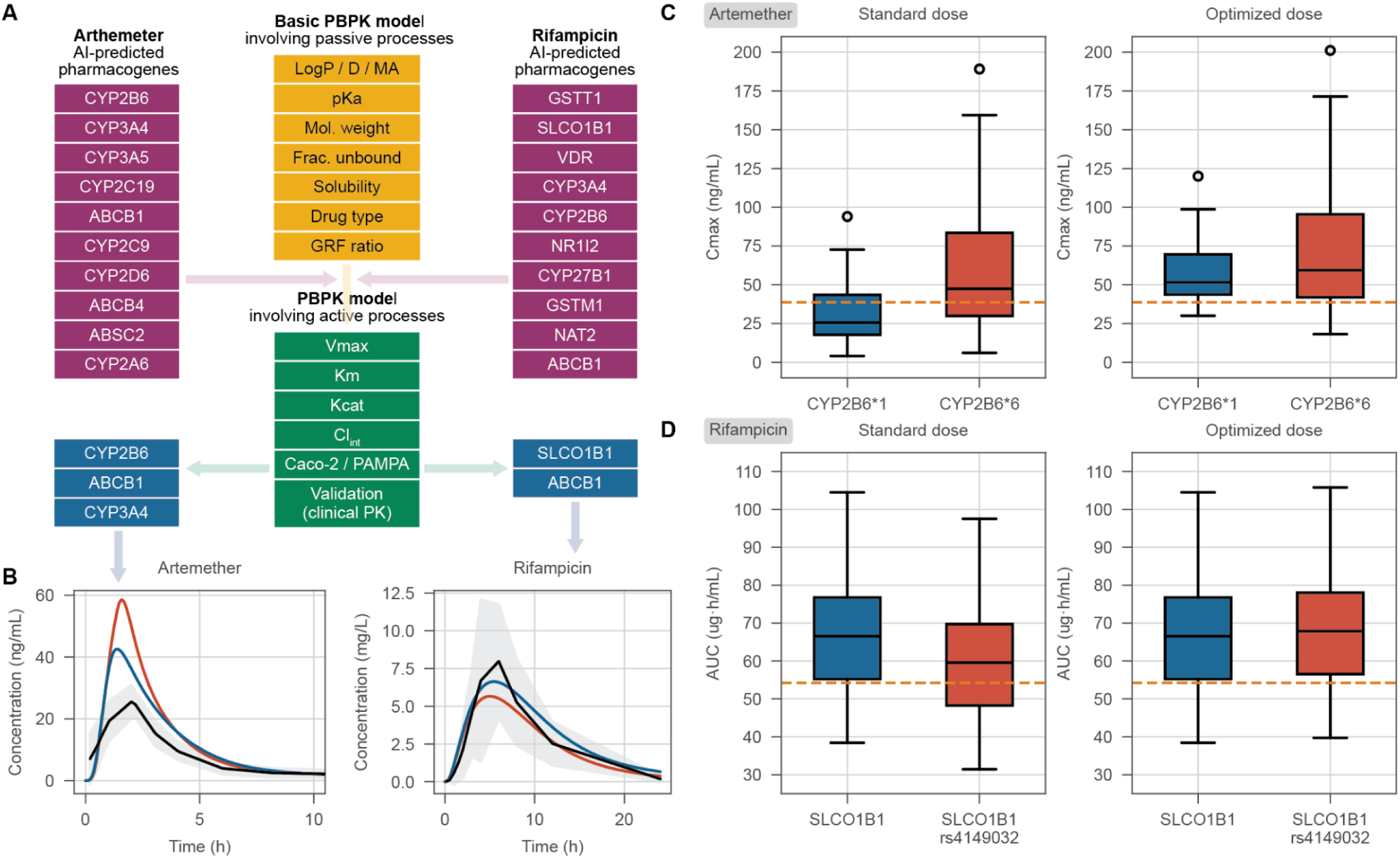
PBPK modelling steps and dose optimisation for artemether and rifampicin. **A.** Building PBPK models for artemether and rifampicin involving integration of AI predicted pharmacogenes and model validation using clinical PK data. **B.** Predicted concentration-time profiles involving active processes of sensitive pharmacogenes with observed clinical PK data. **C** and **D.** NLME modelling, assessment of standard dose of artemether and rifampicin for PK target attainment in African population harbouring enzyme/ transporter variants and Monte Carlo simulation of optimised doses.

### NLME modelling and Monte Carlo simulation for the dose optimisation

Finally, we assessed the above-mentioned genes in the context of a dose optimisation analysis. The NLME modelling of artemether confirmed the CYP2B6*1 and CYP2B6*6 star alleles to be the significant covariates of artemether pharmacokinetics (supplementary Table S6). Virtual patients harbouring CYP2B6*1 and CYP2B6*6 variants were associated with ‘normal’ and ‘high’ plasma drug exposure, respectively. Monte Carlo simulation of artemether standard dose of 80 mg resulted into 41.8% of the population with CYP2B6*1 attaining the Cmax target of 38.6 ng/mL while in the population with CYP2B6*6, 78% attained the target Cmax. The optimum doses necessary for the patient populations with CYP2B6*1 and *6 variants to reach a probability of > 91% target attainment were 152 and 85 mg, respectively (Figure 4A). Similarly, the SLCO1B1 and SLCO1B1 rs4149032 transporter protein variants were confirmed as significant covariates of rifampicin pharmacokinetics. The SLCO1B1 rs4149032 variant was associated with low rifampicin plasma exposure compared with the SLCO1B1 wildtype. Simulation of rifampicin standard dose of 600 mg resulted in 43.5% of the virtual patient population with SLCO1B1 rs4149032 attaining the AUC target of 54.2 µg*h/mL while 91.3% of the virtual patient population with the SLCO1B1 variant attained the AUC target (Figure 4). Monte Carlo simulations showed that the rifampicin dose of 750 mg was optimum in the population of patients with the SLCO1B1 rs4149032 transporter variant as it resulted in over 91% of the population attaining the target AUC.

## Discussion

We have developed an ML/AI and pharmacometrics pipeline for the prioritisation of PGx interactions in low-data scenarios, with the goal to address unmet clinical needs in Africa. Despite recent advances in ML/AI, particularly in foundational LLMs like GPT-4 or Llama 3.1, applications to real-world healthcare issues are still incipient^27^, with very little literature describing AI-based approaches to PGx^28^. In this study, we demonstrated how the systematic exploitation of publicly available PGx data across all genes and disease indications via a biomedical knowledge base, coupled with the use of an LLM for reasoned candidate selection, provides accurate suggestions for the improvement of pharmacometric modelling in African populations. To the best of our knowledge, this is a pioneering effort in the field of PGx, with the potential to set the stage for broader and more in-depth analysis as more data becomes available, potentially beyond the publicly available PharmGKB resource.

As a proof-of-concept, we have focused on malaria and TB, which collectively cause over one million deaths yearly on the African continent^29,30^. While some drugs, such as isoniazid, are relatively well annotated in terms of PGx interactions, other newer drugs, like bedaquiline, lack actionable information that could be crucial to improve the drug dosage in specific populations. Overall, infectious disease drugs are underrepresented in PharmGKB, with only 14.2% of the drugs annotated belonging to this category, and furthermore, when annotated, they present less annotations per drug than non-communicable diseases. We hypothesise these numbers are not due to intrinsic characteristics of infectious disease drugs, but a simple reflection of the status of biomedical research, which is highly skewed towards non-communicable diseases with more relevance to the Global North.

From a methodological standpoint, we propose a pipeline that couples a novel ML/AI component with well-established pharmacometric modelling techniques, namely PBPK modelling, NLME modelling and Monte Carlo simulations. We have restricted the predictions of PGx interactions to ADME genes, which are much more likely to elicit PK effects than non-ADME genes. Using our ML/AI pipeline, we have suggested 10 ADME pharmacogenes for each of the 32 malaria and TB drugs analysed. Unsurprisingly, among the most up ranked genes we identify several members of the CYP450 family, as well as several transporters and carriers. The ML/AI component was, in turn, divided into two steps. In the first, more exploratory step, we recapitulated gene families such as GST that are rarely associated with malaria and TB drugs. In the second, more refined and knowledge-driven step, transporters such as ABCB1, ABCC2 and ABCG2 were further highlighted, as well as frequently-reported enzymes like CYP2C9. Thus, our two-step framework yields both well-supported and relatively unexpected findings, which can be useful depending on the level of annotation of the drug of interest. For the poorly annotated drugs (bedaquiline, rifapentine, and many others) the more conservative (step 2) predictions might be desirable, whereas for better-annotated drugs such as moxifloxacin or primaquine, it may be worth inspecting the more preliminary scores from step 1.

Ultimately, our study aims to unravel PGx associations of relevance to African populations, who encompass the majority of patients affected by malaria^29^ and TB worldwide^30^. To that end, we have labelled variants observed in ADME genes as AFR-abundant if they had a high allele frequency in African populations, and as AFR-specific if, on top of this, the frequency was much higher than in any other biogeographical group. By analysing the 1kGPhg38 population genetics dataset, we add an extra layer of annotation to the ADME genes, highlighting those that harbour a higher number of AFR-abundant or AFR-specific alleles. As population genetics studies of relevance to Africa start becoming more and more available, for example thanks to the H3Africa consortium^31^ or, more broadly, the All of Us Research Program^32^, we expect that our simple annotation approach will gain depth and granularity by accounting for different African subpopulations, which is key given the high genetic diversity existing in the continent.

Nonetheless, our proposed pipeline has caveats and limitations. First and foremost, it is not yet capable of predicting the effects of specific variants, which limits its direct application in designing PGx genotyping panels relevant to Africa. Although we attempted to circumvent this limitation through validation of PBPK models with clinical PK data and NLME modelling, it would have been more desirable to incorporate variant knowledge more expressively earlier in the pipeline, specifically during the knowledge embedding and ML steps. However, the sparseness and poor annotation of variants of PGx interest currently hinders this approach. Additionally, the validation of PBPK models requires either in vitro kinetics parameters of enzyme or transporter protein variants and clinical PK data or combined clinical PK and PGx datasets from which kinetic parameters can be modelled. These studies are not always available in literature, and there is a particular scarcity in studies performed in African patients^33^. This makes it more difficult to confirm whether plausible disposition pathways proposed by ML/AI affect PK - for example, to confirm the involvement of ABCB1 in drugs where its contribution has not previously been reported. The contribution of secondary (minor) disposition pathways is also more difficult to account for - sensitivity analysis during PBPK modelling might suggest that they do not contribute to PK, but they may gain importance if the primary pathways are compromised due to disease, or due to interactions with other drugs, both of which are important factors to consider in malaria and TB combination therapy. Second, our focus in this study has been on PK effects mediated by ADME genes, which only capture a portion of the complexity behind PGx interactions. For example, several of the drug -gene interactions in PharmGKB, and thus in our prediction set, involve the induction or inhibition of disposition of a second drug, rather than the drug to which the PBPK model is applied, making them difficult to capture in our workflow. We will explore the impact of these drug-gene-drug interactions in subsequent work. Exploring beyond ADME genes and investigating efficacy (PD) effects is a natural next step, although coupling this exploration to pharmacometric modelling may be less straightforward. Third, from a technical standpoint, the ML/AI components of our pipeline could be extended and updated as new datasets, methods, and LLMs become available. Our code is modular and designed to accommodate such extensions. Specifically, fine-tuning LLMs to explicitly include a corpus of scientific literature related to PGx could increase the accuracy of predictions and reduce the risk of confabulation by the LLM^34^. Finally, our pipeline has been applied to a privileged set of approved drugs for which a relatively large amount of data is available. To make this pipeline informative for preclinical drug development of new medicines, it will be necessary to carefully determine the minimal molecular information required for reliable predictions and to suggest in-vitro experiments accordingly to gather the necessary data.

### Concluding remarks

We have built a computational pipeline to prioritise pharmacogenes and adjust dosing guidelines for malaria and tuberculosis drugs prescribed in Africa. Our method is designed to be broadly applicable, incorporating a wide array of computational techniques into PGx research, where such methodologies have been historically underexplored. We demonstrated how computational predictions can significantly enhance the understanding of observational drug response data in the African continent, thereby contributing to making PGx research more global and representative. However, we believe that this work is just the first stepping stone toward building an AI-driven, data-rich computational ecosystem for PGx. The next immediate step will be to increase the detail of predictions to the genotype level, identifying not only the pharmacogenes but also the specific variants that may be worth investigating in Africa. To achieve this, it will be necessary to expand the breadth and depth of genomic data available for African populations, and to develop new algorithmic approaches to incorporate variant information into the predictive framework presented here. Additionally, enhancing the method with in-vitro (liver microsome and hepatocyte metabolism kinetics) data and sources such as (liver) biobanks might substantially widen the scope and increase the accuracy of our predictions. We are hopeful that this work will raise awareness of the need for more and better data from Africa, and for Africa, ultimately contributing to greater equity in the PGx field.

## Materials and Methods

### Pharmacogenetics data collection

Raw data was downloaded from PharmGKB in January, 2024 (https://www.pharmgkb.org/). We have collected in a single table all the triplets chemical-gene-variant associations that are recorded as “significant” in PharmGKB or with level of evidence 1,2,3 or 4. The main source files curated include: chemicals.tsv (list of all available drugs), genes.tsv (list of all available genes) and variants.tsv (list of all available variants). SMILES information for each drug has been retrieved either from PharmGKB when available or PubChem otherwise, chemicals without a SMILES annotation have been discarded for downstream analysis. Drug indications have been extracted from DrugBank and manually curated. Variants corresponding to a single haplotype have been curated from the allele_definition_tables available at PharmGKB for each haplotype. The triplet annotations drug-gene-variant come from either the Drug Labels, Clinical Annotations, Clinical Variants or Automated Annotations in the PharmGKB Downloads section. Evidence levels have been used as stated in PharmGKB (https://www.pharmgkb.org/page/clinAnnLevels). Data with evidence level 4 has been discarded for downstream analysis. Data coming from variant Annotations is not associated with a level of evidence, but significance is stated. Only significant associations are kept, and level of Evidence is indicated as 5. Automated annotations, for which no level of evidence was available either, have been given an evidence level of 6. A detailed step by step data curation pipeline can be found in the code repository.

Figure 1 shows several statistics from the PharmGKB dataset, with special focus on communicable disease drugs and ADME genes. For the ‘PGx signatures’ analysis, only drugs with at least 5 annotated genes in PharmGKB were considered. The signature analysis was formulated as a ‘topic modelling’ task whereby drugs were ‘documents’ and pharmacogenes were ‘terms’ in these documents^35^. Occurrence of words was proportional to the number of associated variants (3: 50+ variants, 2: 10-49 variants, 1: 1-9 variants), and a TF-IDF transformation was applied to upweight the least frequent (more informative) genes per drug. Then, a non-negative matrix factorisation (NMF) was applied to decompose the drug-pharmacogenes (DxG) (document-terms) matrix into a drug-signature (Dx10) and signature-gene (10xG) matrix. These matrices were then used to project drugs and genes to a 2D space using tSNE. ‘Importance’ of genes in the signature-gene matrix was simply measured as the sum of scores of each gene across signatures.

Selection of drugs and genes: the study is focused on ADME genes and a subset of approved malaria and tuberculosis drugs (Table 1). The list of genes was based on the PharmADME extended list of ADME genes^8^. Malaria and Tuberculosis drugs were manually selected to include all current first line drugs used for treatment, as well as additional structurally diverse drugs useful in the management of these diseases.

**Table 1.**
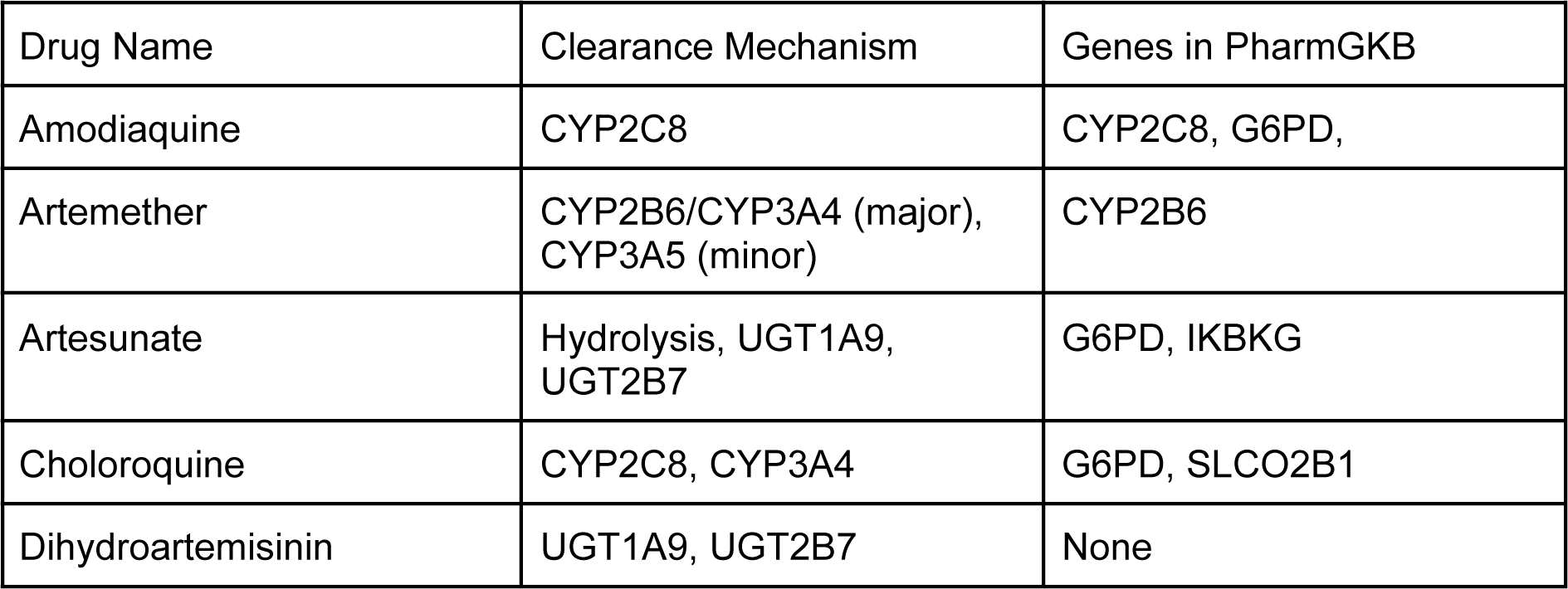
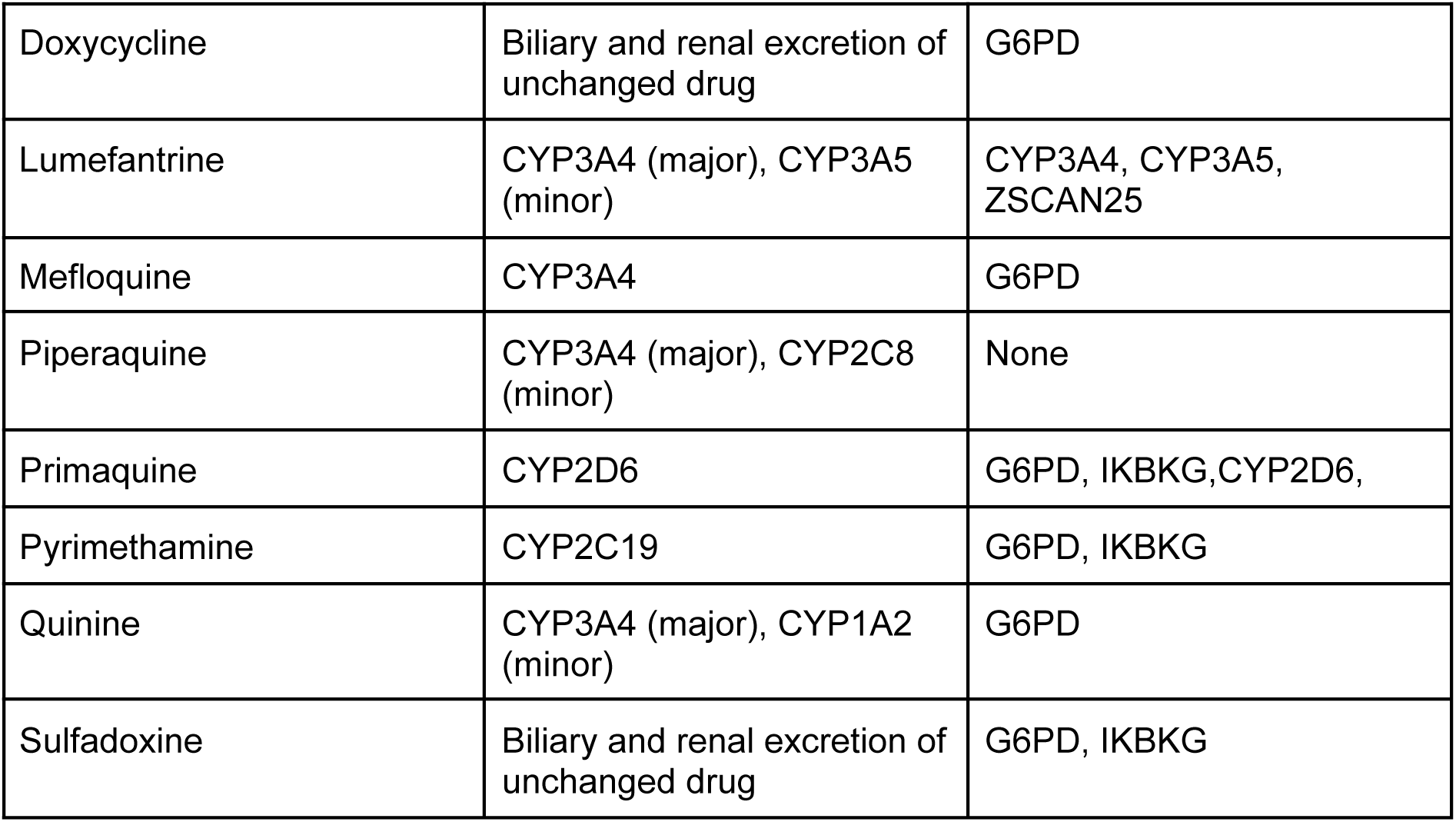
Malaria drugs analysed in this study, ordered alphabetically by name.

**Table 2.**
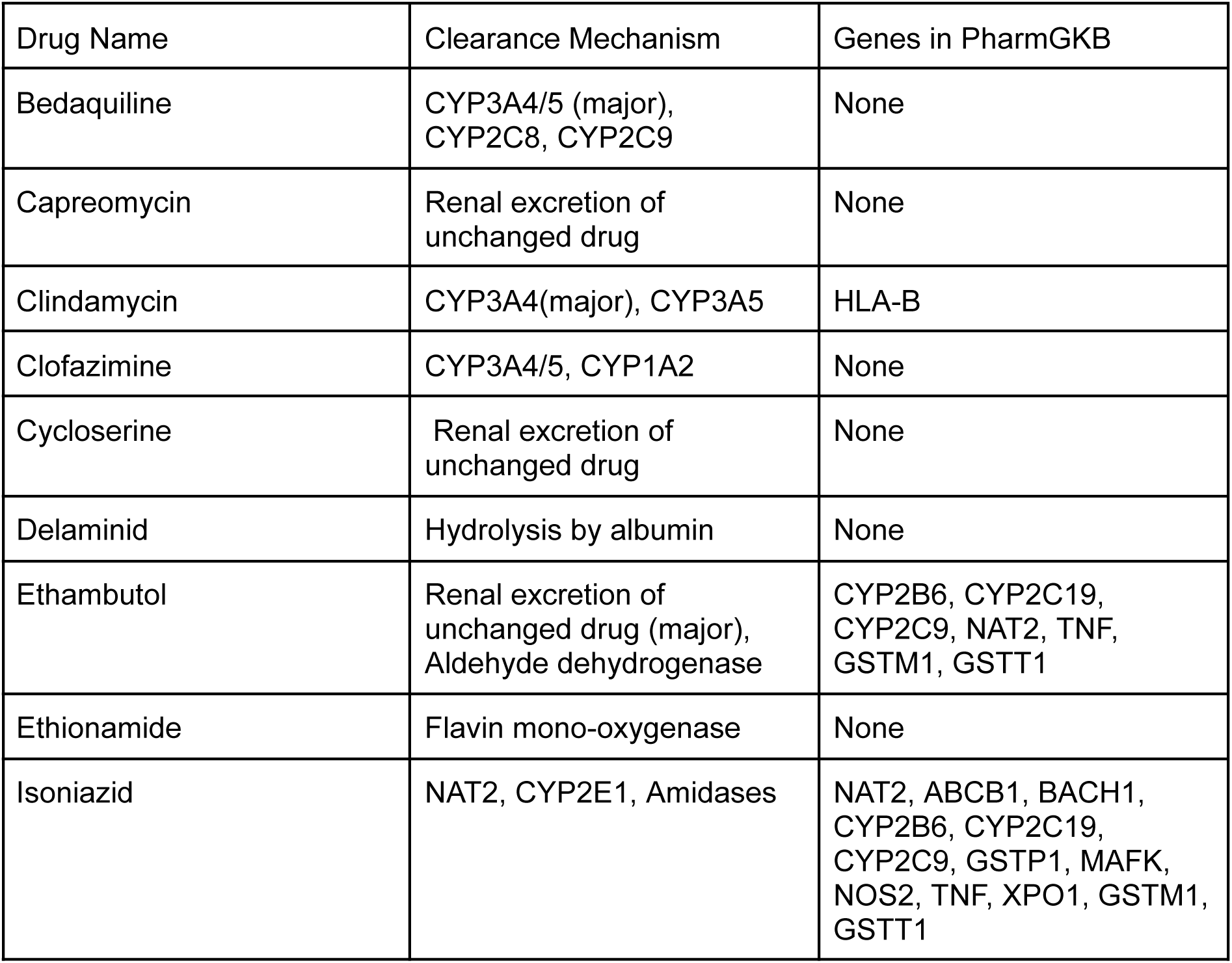
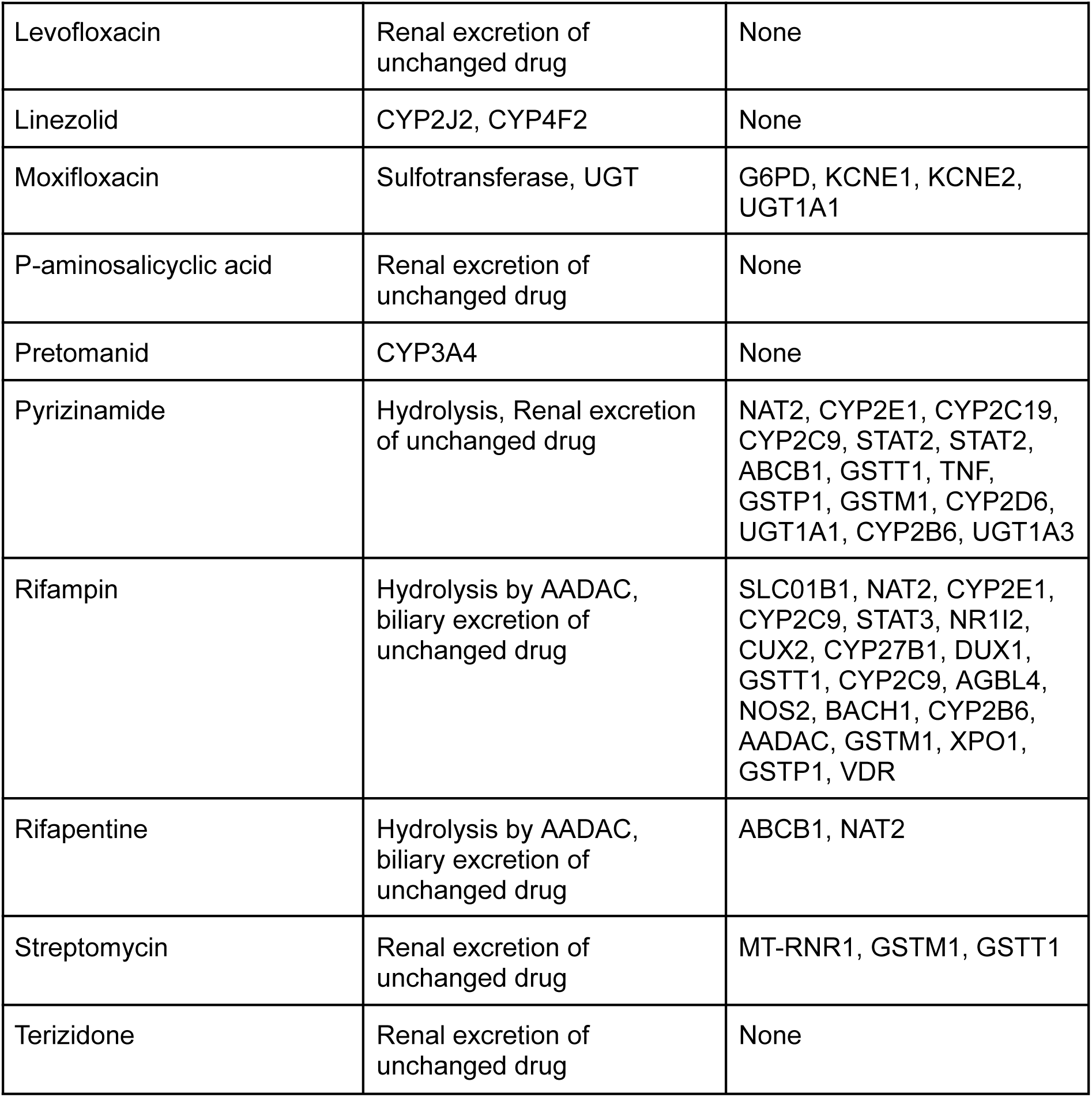
Tuberculosis drugs analysed in this study, ordered alphabetically by name.

### Population genetics data annotation

The 1kGPhg38 dataset for chromosomes 1 to 22 and X was downloaded from the International Genome Sample Resource (IGSR) from the collection named ‘1000 Genomes on GRCh38^36^. A coordinates file of the GENCODE V43 track (genome assembly version GRCh38.p13; ^37^) was obtained from the UCSC Table Browser and the BEDtools software^37,38^ was used to subset those variants found within the GENCODE V43 defined regions (i.e., variants included within exons and introns as well as 5’ and 3’UTRs plus a 200bp downstream and upstream window). Variants in each chromosome were annotated in parallel using the SnpEff/SnpSift set of tools ^22^. Additionally, a total of 69,299 canonical transcripts (Ensembl IDs) from the Human genes GRCh38.p14 dataset obtained from BioMart were used in the annotation process ^39^. The number of initial transcripts in the 100GPhg38 dataset was 78,229,655, all of which were biallellic SNVs. Next, variants were merged and only SNVs within protein coding transcripts were considered for downstream analyses.

### Africa-abundant and Africa-specific variants

African variants are defined from the point of view of their abundance in African populations and their specificity to African populations based on the allele frequency (AF) of a given variant over the populations in the data set. Briefly, an AFR-abundant variant was defined as a variant having an AF over 0.20 in African populations regardless of its AF in the remaining populations, and an AFR-specific variant was defined as a variant within the set of AFR-abundant variants overrepresented at least x8 with regards to the remaining populations.For the determination of an AFR-abundant optimal threshold, precision and recall metrics were calculated for a range of African AFs from 0 to 1 using 0.05 steps. For the determination of an AFR-specific threshold, instead, precision and recall metrics were calculated for a range of African overrepresentation (or specificity) values from 1 to 50. Truly African variants for the calculation of precision and recall were obtained from protein coding variants identified as AFR-specific in the work of Fedorova et al, 2022^10^.

### Orthogonal gene and drug descriptors

Gene descriptors were obtained mainly from the Bioteque resource v1^12^ (for details, see Table S1). In addition, we obtained precalculated protein sequence embeddings from UniProt (ProtT5) and we calculated ESM1b embeddings. Drug descriptors were calculated primarily using the Ersilia Model Hub, which wraps descriptor calculators of multiple types including bioactivity profiles from the Chemical Checker^17^, physicochemical properties, and ADMET calculations^16^ (for details, see Table S3). Robust scaling was applied to normalise each column in these datasets. We also fetched drug descriptors from the Bioteque v1 as detailed in Table S2.

### Embedding-based pharmacogene prioritisation

We applied a supervised learning approach to train a drug-pharmacogene prioritisation model based on drug and gene descriptors. From PharmGKB data, three ground-truth drug-gene pair datasets were derived. First, a large one containing all drugs and all genes, and any pharmacogenetic interaction (model A). Second, a reduced one containing all drugs and all genes, but only considering Metabolism/PK interactions (model B). Finally, a smaller one containing only ADME genes with PK interactions (model C). We therefore trained three classification models using the same pipeline.

For each set of positive data, we sampled random unlabelled drug-gene pairs preserving drug and gene frequencies. The positive:negative ratio was 1:10. Then, we formulated the training as a binary classification task (1: known PGx interaction, 0: no PGx interaction) taking as features the drug and gene descriptors. Globally, this encompassed an ensemble of individual classifiers corresponding to any combination of drug and gene descriptor types. To harmonise the procedure, all descriptors were first reduced to their first 100 PCA components. Then, the drug and gene vectors were concatenated and subsequently reduced to 50 components using linear-optimal low-rank projection (LOLP^40^). As a supervised learning algorithm, we used the LGBM zero-shot base classifier as provided by the AutoML tool FLAML^41^. To obtain a single prediction across the ensemble, we calculated the weighted average of all individual predictions, with weights directly proportional to the individual AUROC performance within the range 0.5-1. Of note, not all drugs and genes have descriptors of all types. The weighted average was only calculated on the applicable individual classifiers, and a ‘support’ measure was appended to the output quantifying the number of individual models participating in the ensemble-based prediction. Models were evaluated with a stratified 5-fold cross-validation scheme, using AUROC as the main performance metric.

The trained models were used to explore exhaustively all possible drug-gene pairs. Predictions from models A, B and C were aggregated in a consensus score using a simple average, and an empirical z-score was calculated using all predictions as background. Here, we focused on the set of 32 malaria and TB drugs. For each drug, we ranked ADME genes according to their z-score. The top 50 ADME genes per drug were kept for further analysis (Ranked List 1).

### LLM-based pharmacogene re-ranking

Ranked List 1 was then submitted to a re-ranking procedure based on LLMs. GPT-4 was used as the main LLM throughout the pipeline. As a first step, we aimed at obtaining succinct and structured TLDR reports for drugs and genes. For drug TLDRs, the LLM was presented with the DrugBank v5 information, and the LLM prompt was engineered to provide a drug summary, a paragraph describing targets, enzymes, transporters and carriers, and a paragraph on the known pharmacogenetics of the drug. Similarly, for gene TLDRs, the prompt was engineered to provide a TLDR consisting of a short summary of the gene, including generic knowledge of its function, a brief summary of the associated diseases, phenotypes, pathways, and drugs (if any), and a paragraph about pharmacogenetic knowledge of the gene.

These drug and gene TLDR summaries were used as context for the re-ranking LLM query. In this case, for each drug, the top 50 genes from Ranked List 1 were provided, along with their summaries. In addition, known associations in PharmGKB were given as context. The prompt was engineered such that the LLM was encouraged to make inferences in the ‘role’ of a pharmacogenetics expert, based on knowledge about similar drugs and genes, mechanisms of action, etc. Table S4 shows a simplified version of this and other prompts. The LLM was asked to provide a list of top 10 pharmacogenes per drug (Ranked List 2) based on the original ranking of 50. The query was performed three times to minimise the chance of confabulation. Importantly, the LLM was also asked to provide a brief explanation for each drug-pharmacogene association.

### PBPK-based sensitivity analysis

The enzymes and/or transporter proteins encoded by the top 10 pharmacogenes for rifampicin, bedaquiline, artemether and amodiaquine were separately inputted into the PBPK model implemented in PK-Sim^Ⓡ^ v11.3. These models were previously validated using pharmacokinetic data from published clinical studies performed in cohorts from the African populations. In order to perform sensitivity analyses, each enzyme or transporter protein was assigned a random initial value for *in vitro* kinetic parameters such maximum rate of reaction (V_max_), Michaelis constant (Km) and intrinsic clearance (CL_int_). This step was necessary for the systematic assessment of how variations or uncertainties in model parameters affect model prediction. Model parameter sensitivity was calculated according to Eq. (1),

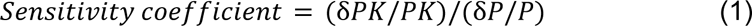

where, PK is the initial value of the pharmacokinetic parameter such as area under the curve (AUC), maximum concentration (C_max_), apparent clearance (Cl/F), apparent volume of distribution (Vd/F), time at which concentration reaches its maximum (T_max_), half-life (T_Half_) or mean residence time (MRT). δPK is the change of the pharmacokinetic parameters from initial values. P is the initial value of the evaluated input parameter. δP is the change of evaluated input parameters from initial value, respectively. A sensitivity coefficient of 1.0 indicates that 10% change of an evaluated input parameter causes 10% change of the predicted pharmacokinetic parameter. Therefore, we set the sensitivity coefficient of 1 as the minimum criterion of considering a pharmacogene (enzyme or transporter protein) as ‘sensitive’ in the PBPK model. The sensitive and known pharmacogenes from literature together with their variants were retained in the PBPK model for further parameter identification. The predicted concentrations were compared with the observed as a way of model validation or confirmation.

### NLME modelling

The PBPK predicted concentration-time profiles each drug stratified by enzyme or transporter protein variant were preprocessed in R for NLME modelling. The datasets of virtual African patients included enzyme or transporter variants as covariates. The Monolix suite 2024R1 (https://lixoft.com/products/monolix/) was used to perform NLME and covariate modelling and determine whether covariates were significantly affecting the pharmacokinetics of the drugs. This was followed by Monte Carlo simulation to assess the current standardised dose and determined optimum dose in African patient populations harbouring certain enzyme or transporter variants.

## Data availability

All PGx data used in the study has been curated from the publicly available resource PharmGKB, accessed in January, 2024. Curated data is available in the https://github.com/ersilia-os/pharmacogx-embeddings. Data related to genomic variants has been curated from the 1000 Genomes project and is available in https://github.com/ersilia-os/pharmacogx-arsa.

## Code availability

Code for the data curation and training of the PGx classifier can be found in https://github.com/ersilia-os/pharmacogx-embeddings. Code for the annotation of genomic variants based on the 1kGPhg38 can be found in https://github.com/ersilia-os/pharmacogx-arsa. A simple app showing the final results of the publication is available at https://github.com/ersilia-os/pharmacogx-app and is demoed at https://pharmacogx-embeddings.streamlit.app. All code is licensed under a GPLv3 License. Note that these tools are intended for research purposes only and should not be used in clinical practice.

## Acknowledgements

Research reported in this publication was supported by the South African Medical research Council (SAMRC) with funds received from Novartis and GSK R&D for Project Africa GRADIENT (Grant # GSKNVS1/202101/004). The authors are grateful for helpful discussions with Dr Adolfo Garcia-Perez (GSK) and Dr Pablo Castaneda-Casado (GSK) which helped define the strategy in this project.

## Supplementary Information

**Table S1.** Bioteque gene embeddings.

| Metapath | Subset | Description |
| --- | --- | --- |
| GEN-ass-DIS | Open Targets | Disease-gene associations from Open Targets |
| GEN-ass-DIS | DisGeNeT (curated) | Disease-gene associations from DisGeNeT (curated) |
| GEN-has-MFN | GO Molecular Function | Gene Ontology molecular functions as reported in GOA |
| GEN-has-CMP | Jenssen Compartments | Cellular localization data for genes |
| GEN-ass-PWY | Reactome | Pathway annotations from Reactome |
| GEN-ppi-GEN | STRING | Functional protein-protein interactions (PPIs) from STRING |
| GEN-ppi-GEN | HURI (union) | Human Reference Interactome (HuRI) PPIs (yeast two-hybrid dataset) |
| GEN-ppi-GEN | HI Y2H (union) | PPIs from the HI Y2H dataset |
| GEN-pab-TIS | HPA Proteome | Human Protein Atlas abundance in tissues |
| GEN-pdf-TIS | HPA Proteome | Human Protein Atlas deficiency in tissues |
| GEN-has-DOM | InterPro | Protein-domain associations based on InterPro |
| GEN-upr-CLL | GDSC1000 mRNA | Upregulated genes in the GDSC cell line panel |
| GEN-cex-GEN | CoexpressDB | Gene-gene coexpression from CoexpressDB |

**Table S2.** Bioteque drug embeddings.

| Metapath | Subset | Description |
| --- | --- | --- |
| CPD-int-GEN | Curated Targets | Drug-protein interactions from DrugBank, RepoHub and PharmacoDB |
| CPD-int-GEN | DrugBank | Drug-protein interactions from DrugBank |
| CPD-int-GEN | DrugBank (PD) | Drug-target interactions from DrugBank |
| CPD-int-GEN | DrugBank (PK) | Drug-enzyme, drug-transporter and drug-carrier interactions from DrugBank |
| CPD-int-GEN | DrugCentral | Drug-target interactions from DrugCentral |
| CPD-int-GEN | PharmacoDB Targets | Drug targets from the cancer cell line panels resource PharmacoDB |
| CPD-int-GEN | RepoHub | Drug targets from the Drug Repurposing Hub |
| CPD-cau-DIS | CTD | Causative compound-disease associations from CTD |
| CPD-cau-DIS | OFFSIDES & SIDER | Drug side effects from OFFSIDES and SIDER |

**Table S3.**
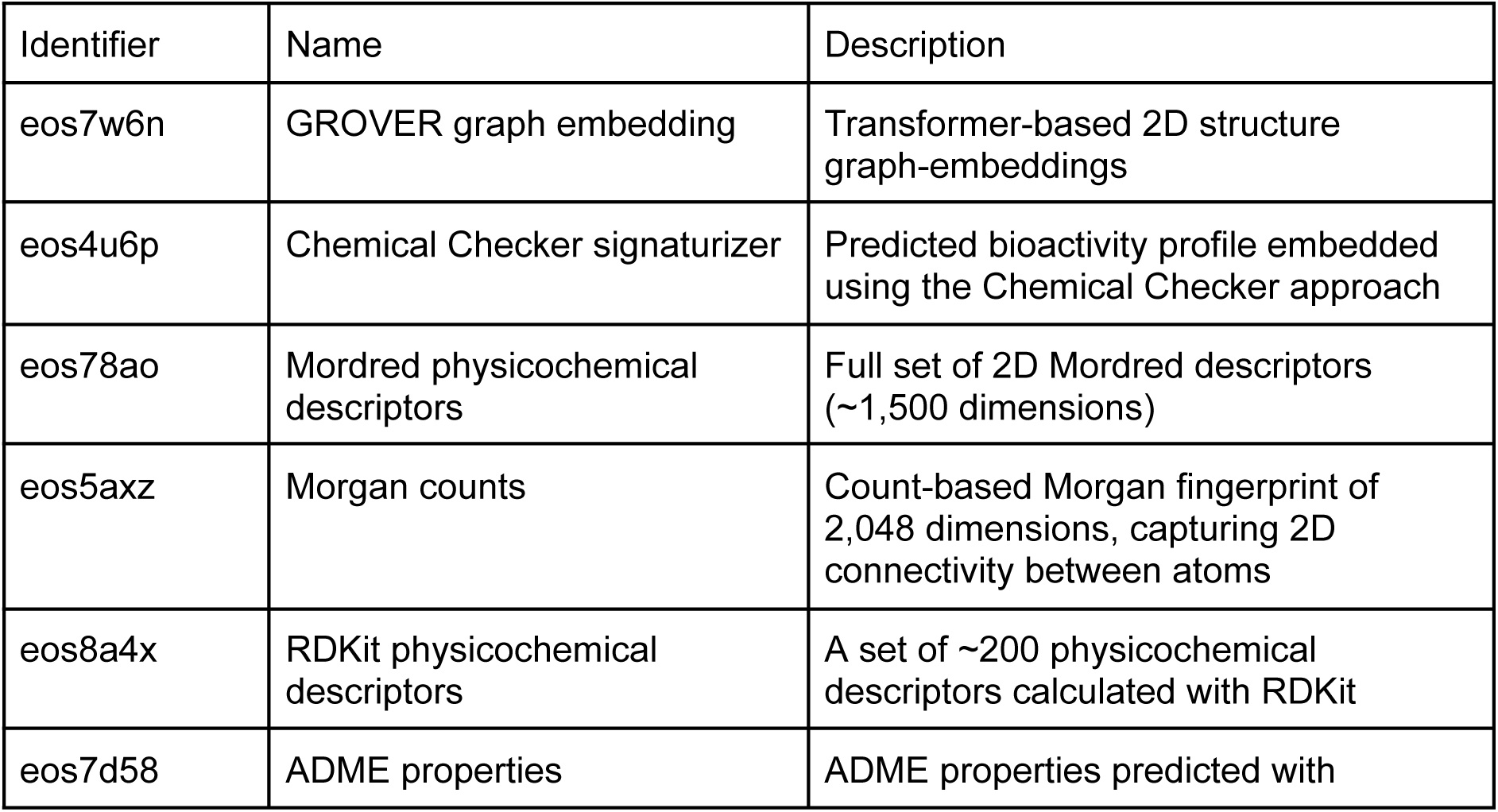
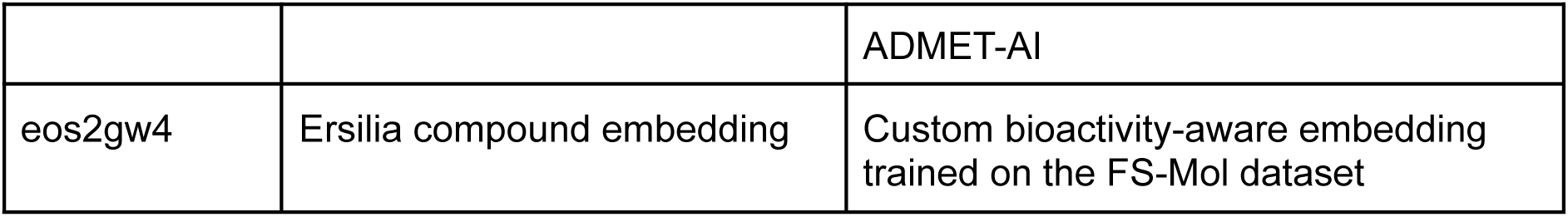
Ersilia Model Hub drug descriptors.

| Identifier | Name | Description |
| --- | --- | --- |
| eos7w6n | GROVER graph embedding | Transformer-based 2D structure graph-embeddings |
| eos4u6p | Chemical Checker signaturizer | Predicted bioactivity profile embedded using the Chemical Checker approach |
| eos78ao | Mordred physicochemical descriptors | Full set of 2D Mordred descriptors (~1,500 dimensions) |
| eos5axz | Morgan counts | Count-based Morgan fingerprint of 2,048 dimensions, capturing 2D connectivity between atoms |
| eos8a4x | RDKit physicochemical descriptors | A set of ~200 physicochemical descriptors calculated with RDKit |
| eos7d58 | ADME properties | ADME properties predicted with |
|  |  | ADMET-AI |
| eos2gw4 | Ersilia compound embedding | Custom bioactivity-aware embedding trained on the FS-Mol dataset |

**Table S4.** LLM prompts.

| Concept | LLM prompt (slightly adapted and shortened version, excluding formatting details) |
| --- | --- |
| Drug TLDR | <u>System prompt:</u><br>You are a pharmacogenetics expert. You are asked to provide a summary of a given drug. You'll be provided with two pieces of information: (a) a name of a drug and its synonyms, and (b) an extract of data from DrugBank, containing free text data as well as lists targets, enzymes, transporters, etc. You should use the information in (b) but you should also use your own knowledge to provide a summary of the drug. Do not restrict yourself to DrugBank information, especially for pharmacogenetics. Other useful resources are PharmGKB, FDA drug labels, and the scientific literature. Feel free to infer pharmacogenetic interactions, but clarify if you are making an inference.<br>Your summary should be structured as follows: (1) A summary paragraph of the drug, including its name, therapeutic class, and general knowledge about its pharmacokinetics, pharmacodynamics and metabolism. (2) A summary of the drug's targets, enzymes, transporters and carriers. (3) A paragraph on drug's pharmacogenetics, including any known association to pharmacogenes.<br>Be as succinct as possible. Your response should not exceed 500 words. Strictly limit yourself to 3 paragraphs.<br><u>User prompt:</u><br>Provide a summary of the drug given the following information: (a) Drug name: [...], DrugBank information: [...]. |
| Gene TLDR | <u>System prompt:</u><br>You are a pharmacogenetics expert. You are asked to provide a summary of a given gene. You'll be provided with a gene name or symbol. You should use your own knowledge to provide a summary of the gene. Useful resources include PharmGKB, UniProt, Gene Cards, and the scientific literature.<br>Your summary should be structured as follows: (1) A summary paragraph of the gene, including its official symbol, official name, and general knowledge about its function and expression. (2) A |
|  | summary of the gene's drugs, diseases, phenotypes, and pathways. (3) A paragraph on the gene's pharmacogenetics, including any known pharmacogenetic associations with drugs. Mention as many drugs as possible.<br>Be as succinct as possible. Your response should be at most 500 words long. Do not use full gene names. Use the official gene symbols.<br><u>User prompt:</u><br>Provide a summary of the gene: [...] |
| Drug paragraph | <u>System prompt:</u><br>You are a pharmacogenetics expert. You'll be asked to provide a short summary of a given drug's pharmacogenetic interactions profile. You will be provided with a drug name and a summary of the knowledge on the drug. In addition, you may be provided with a list of genes that are known to interact with the drug.<br>You should provide a summary of the drug's pharmacogenetic interactions profile. Your answer should be between 100 and 200 words, in one paragraph. Mention all genes provided in the list and try to offer an explanation for each of them. You should try to distinguish between pharmacokinetics and pharmacodynamics.<br><u>User prompt:</u><br>Summarise the following text in one or two sentences. Focus on the genes and their interactions with the drug, and on the explanation: [Drug TLDR]<br>Pharmacogenetic knowledge: (a) Pharmacokinetics and ADME genes: [...]. (b) Pharmacokinetics and other genes: [...]. (c) Possibly non-pharmacokinetics and ADME genes: [...]. (d) Possibly non-pharmacokinetics and other genes: [...] |
| Gene paragraph | <u>System prompt:</u><br>You are a pharmacogenetics expert. You'll be asked to provide a short summary of a given gene's pharmacogenetic interactions profile with drugs. You will be provided with a gene symbol and a short summary for the gene. In addition, you may be provided with a list of drugs that are known to interact pharmacogenetically with the gene.<br>You provide a summary of the genes's pharmacogenetic interactions profile. Your answer should be between 100 and 200 words. Mention all drugs provided in the list and try to explain each of them. You should try to distinguish between pharmacokinetics and pharmacodynamics.<br><u>User prompt:</u> |
|  | Summarise the following text in one or two sentences. Focus on the drugs and their interactions with the gene, and on the explanation: [Gene TLDR]<br>Pharmacogenetic knowledge: (a) Pharmacokinetics: [...]. (b) Possibly non-pharmacokinetics: [...] |
| Drug-pharmacogene reranking | <u>System prompt:</u><br>You are a pharmacogenetics expert. Your goal is to identify pharmacogenetic drug-gene pairs. The user will specify a drug of interest. You need to rank a set of candidate genes according to their likelihood of being pharmacogenetically related to the drug. Prioritise pharmacokinetic interactions, such as those affecting drug metabolism, transport, and excretion.<br>The user will provide a pre-ranked list of 50 genes, along with a z-score. Z-scores above 1.96 are significant. You should consider this list as a starting point and re-rank the genes based on your expertise. Up-rank genes with a known pharmacogenetic relationship with the drug. Do not include genes that are not in the pre-ranked list.<br>The user may also provide auxiliary information about the drug and the genes. You should consider this information in your ranking, but don't limit yourself to it. Do not focus only on known associations. Use your expertise to infer new pharmacogenetic relationships. Make logical and mechanistic reasoning based on gene function, gene expression, drug mechanism of action, pharmacokinetics, etc. For example, consider known pharmacogenetic relationships of similar drugs and similar genes in your reasoning.<br>You should return a ranked list of genes. The list should be sorted in descending order of likelihood. Give me only the top 10 genes.<br>For each gene, offer an explanation of why you think there is a pharmacogenetic relationship with the given drug. This explanation should be 500 words long. Be as detailed as possible. The explanation should be detailed enough to convince a biomedicine expert that the gene is likely to be pharmacogenetically related to the drug.<br><u>User prompt:</u><br>My drug of interest is: [...]<br>These are the candidate genes for this drug, tentatively ranked by the Z-score: [Ranked List 1]<br>In addition, consider that the following pharmacogenetic interactions are already known for this drug, according to PharmGKB: [...]<br>Below is auxiliary information about the given drug: [Drug |
|  | paragraph]<br>Below is auxiliary information about the genes in the pre-ranked list: [Gene paragraph] |

**Table S5:** A list of antimalarial and antituberculosis drugs with corresponding percentage interindividual (IIV) variability in apparent clearance (CL/F) and apparent volume of distribution (Vd/F).

| Drug | IIV, CI/F (%) | IIV, Vd/F (%) |
| --- | --- | --- |
| Artemether | 28 - 99 | 20.5 |
| Lumefantrine | 14.8 - 38 | 15.2 - 48.7 |
| Quinine | 40 - 73.6 | 44.5 - 65.3 |
| Artesunate | 43.4 | 76.7 - 77.7 |
| Sulfadoxine | 34 - 65.7 | 11.2 - 16 |
| Pyrimethamine | 35.6 - 84 | 16.7 - 106 |
| Amodiaquine | 32 | 53 |
| Dihydroartemisinin | 20.7 - 90.4 | 34.7 - 60 |
| Piperaquine | 45 | 65 |
| Doxycycline | - | - |
| Clindamycin | - | - |
| Chloroquine | 30 | 62 |
| Primaquine | 65.3 | 82.9 |
| Mefloquine | 12 - 48 | 19 - 90 |
| Proguanil | 22.6 | 17 |
| Atovaquone | 68 | 46.8 |
| Tafenoquine | - | - |
| Streptomycin | - | - |
| Rifampicin | 5.5 - 30 | 19 - 49 |
| Isoniazid | 18 - 93 | 16 |
| Pyrazinamide | 20 - 61.2 | 16 - 39.4 |
| Ethambutol | 20 |  |
| Ethionamide | 120 | 168 |
| Rifapentine | 22 | 16 |
| Moxifloxacin | 13 - 33 | - |
| Levofloxacin | 15.2 | - |
| Bedaquiline | 39 | 41 |
| Linezolid | 37 | 32 |
| Clofazimine | 25.6 | 54.6 |
| Cycloserine | 32.7 | - |
| Terizidone | 64 | 16 |
| Delamanid | 20 | 26 |
| P-aminosalicylic acid | 47.5 - 177 | 74.8 |
| Pretomanid | 19 | - |
| Capreomycin | - | 123 |
| Sutezolid | - | - |

**Table S6:** Physiologically based pharmacokinetic model parameters for artemether and rifampicin in the African population. Kcat = in vitro Vmax per recombinant enzyme. a = Parameter optimised on clinical pharmacokinetics data. b = Assumed the drug is passively filtered from kidney blood plasma into urine. NA = Not applicable.

| Physicochemical and ADME properties | Artemether | Rifampicin |
| --- | --- | --- |
| Molecular weight (g/mol) | 298.38 | 822.9 |
| Log D/ P | 3.28 <sup>1,5</sup> | 2.7 <sup>2</sup> |
| Fraction unbound | 0.046<sup>1</sup> | 0.116<sup>3</sup> |
| Compound type | Neutral<sup>1</sup> | Ampholyte<sup>3</sup> |
| pka | NA | 1.7 and 7.9<sup>3</sup> |
| Solubility (mg/L) | 166<sup>5</sup> | 1400<sup>6</sup> |
| Caco-2 (cm/s x 10<sup>-6</sup>) | 42.7<sup>1</sup> | 5.79<sup>4</sup> |
| AADAC |  |  |
| Kcat (1/min) | - | 6.5<sup>a,7,8</sup> |
| ABCB1 |  |  |
| Kcat (1/min) | - | 47.83<sup>a,7,8</sup> |
| SLCO1B1 |  |  |
| Kcat (1/min) | - | 0.001<sup>a,7,8</sup> |
| SLCO1B1 rs4149032 |  |  |
| Kcat (1/min) | - | 2.87<sup>a,7,8</sup> |
| CYP2B6*1 |  |  |
| Km (μM) | 3.1<sup>9</sup> | - |
| Vmax (pmol/min/pmol) | 36<sup>9</sup> | - |
| Kcat (1/min) | 51.44<sup>a,10</sup> | - |
| CYP2B6*6 |  |  |
| Km (μM) | 6.72<sup>9</sup> | - |
| Vmax (pmol/min/pmol) | 150<sup>9</sup> | - |
| Kcat (1/min) | 17.9<sup>a,10</sup> | - |
| CYP3A4 |  |  |
| Km ($\mu\text{M}$) | 8.24<sup>9</sup> | - |
| Vmax (pmol/min/pmol) | 12.3<sup>9</sup> | - |
| Kcat (1/min) | 0.000578<sup>a,10</sup> | - |
| GFR fraction | 1<sup>b</sup> | 1<sup>b</sup> |

**Figure S1.**
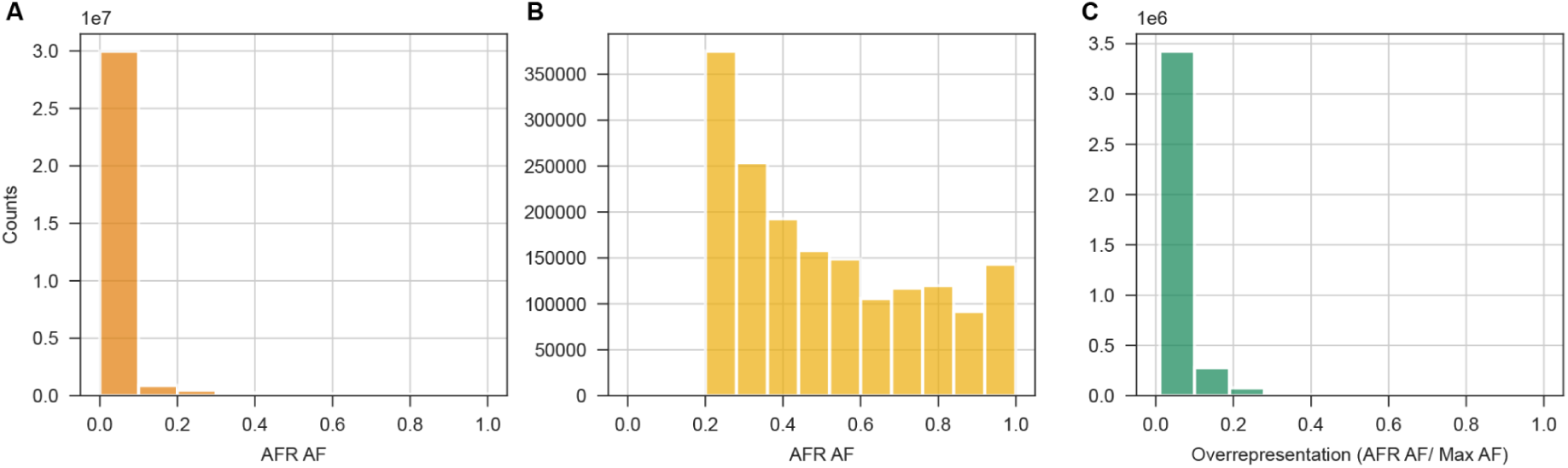
Distribution of African Allele Frequencies. **A.** Allele Frequency distribution of SNVs in African populations (AFR AF) in the filtered 1kGPhg38 dataset containing 32,577,573 SNVs annotations. **B.** Distribution of the African Allele Frequency in the 1,734,675 set of AFR-abundant variants. **C.** Overrepresentation distribution of African specific variants vs other populations using all the populations found within the AFR population group.

**Figure S2.**
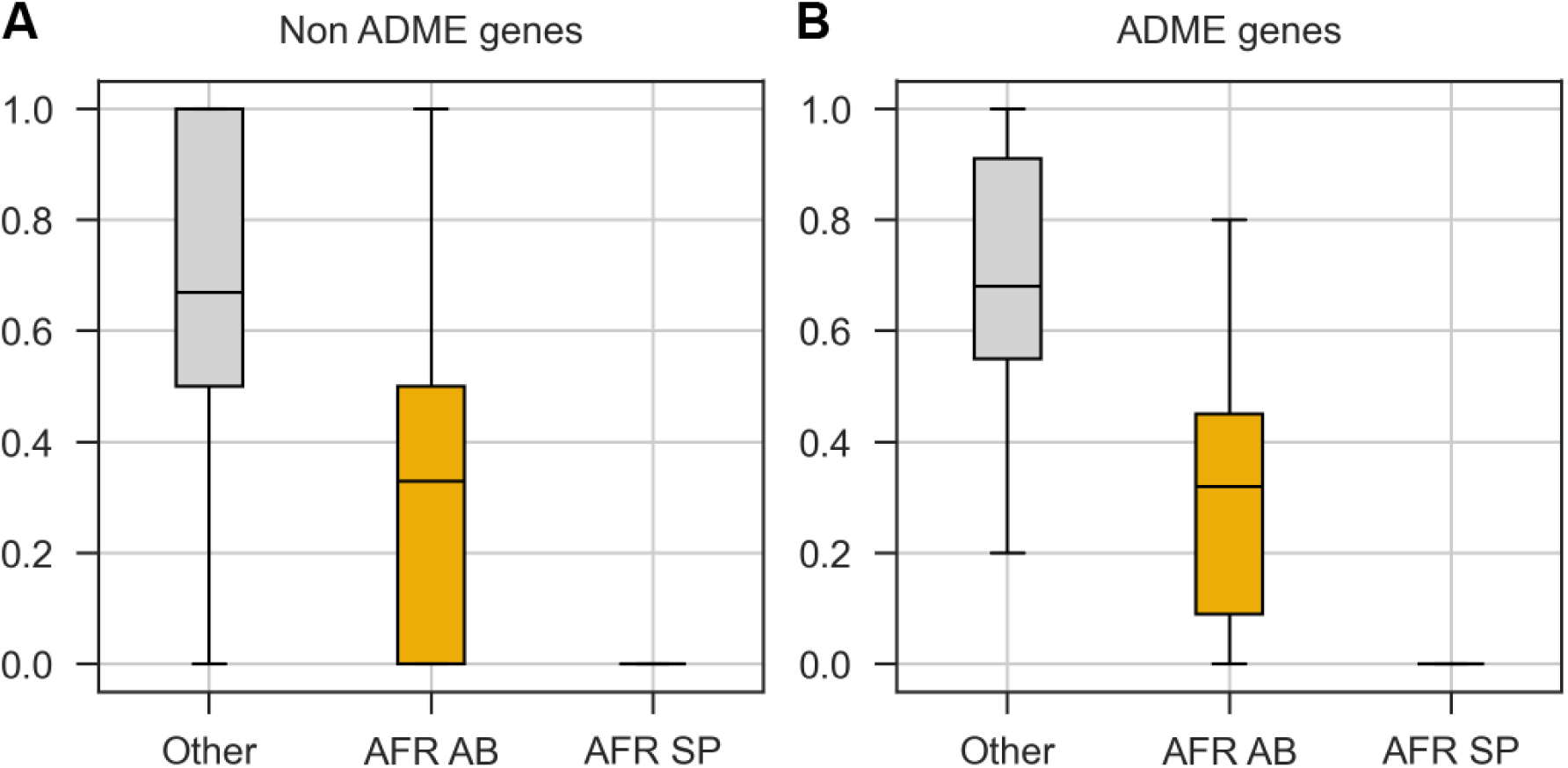
Proportion of African Abundant and African Specific variants in PharmGKB annotated genes. **A.** Frequency of AFR-abundant, AFR-specific and other variants annotated in PharmGKB for non-ADME genes (1195). **B.** Frequency of AFR-abundant, AFR-specific and other variants annotated in PharmGKB for ADME genes (167).

